# A Quantitative Trait Locus for Reduced Microglial *APOE* Expression Associates with Reduced Cerebral Amyloid Angiopathy

**DOI:** 10.1101/2025.03.10.25323519

**Authors:** Michael E. Belloy, Jonathan Graff-Radford, Michael D. Greicius

**Affiliations:** NeuroGenomics and Informatics Center, Washington University School of Medicine, St.Louis, MO, USA; Department of Neurology, Washington University School of Medicine, St.Louis, MO, USA; Departments of Neurology, Mayo Clinic, Rochester, MN, USA; Department of Neurology and Neurological Sciences, Stanford University School of Medicine, Stanford, CA, USA

## Abstract

The *Apolipoprotein E (APOE)* e4 and e2 alleles are respectively the most risk increasing and risk decreasing, common genetic risk factors for Alzheimer’s disease (AD)^1,2^. They strongly affect Aβ burden in the brain parenchyma^1^, a core hallmark of AD, but also at the level of the brain vasculature, i.e. cerebral amyloid angiopathy (CAA)^1,3^, which in turn relates to increased risk for amyloid-related imaging abnormalities (ARIA) in *APOE*\*4 carriers when receiving anti-Aβ antibody treatments^4^. This makes *APOE* a highly pursued AD drug target. A crucial question in the field is whether it would be beneficial to either increase or decrease *APOE* (particularly *APOE*\*4) levels^5^. The answer from rodent work appears to converge on “decreasing *APOE* levels”^5–7^, with initial human studies supporting this^5,8,9^. Human genetic evidence however remains scarce and new insights are crucially needed to support clinical translation. Shade et al. 2024 conducted the largest to date genome-wide association study (GWAS) of various neuropathological traits, identifying a variant protective of CAA in the *APOE* locus independent of *APOE*\*4 and *APOE*\*2 genotypes^10^. Downstream analyses suggested this signal links to the nearby *APOC2* gene through local effects on methylation. We applaud the authors on their timely, relevant, and well-conducted study. Here, we extend on these findings, highlighting there is compelling evidence that their genetic signal for reduced CAA relates to an effect on reduced microglial *APOE* expression, which would importantly support the evidence in favor of “decreasing *APOE* levels” and further herald this promising therapeutic avenue, not just for AD, but also for CAA. We additionally provide complimentary results regarding this locus’ association with CAA and AD risk from analyses that we conducted parallel to Shade et al. 2024.

## Introduction

Shade et al performed GWAS of neuropathological traits across the National Alzheimer’s Coordinating Center (NACC), the Religious Orders Study and Rush Memory and Aging Project (ROSMAP), and the Adult Changes in Thought (ACT) study. Their CAA GWAS identified rs7247551 at the *APOE* locus after conditioning on *APOE*\*4 and *APOE*\*2 genotypes (**Table.1**). They further conducted genetic colocalization analyses with expression quantitative trait locus (e)(QTL) data in 48 tissues (including brain) from GTEx and in brain dorsolateral prefrontal cortex (DLPFC) from ROSMAP, as well as methylation (m)QTL data in DLPFC from ROSMAP. While their CAA signal colocalized with *APOC2* eQTLs in multiple brain tissues in GTEx, it did not in ROSMAP. In fact, their top variant, rs7247551, had opposite effects on *APOC2* expression in brain tissues across GTEx and ROSMAP (and was only nominally associated in the latter, P=0.00013). There was not an *APOE* eQTL effect in brain for rs7247551 in either dataset. They did observe colocalization with DLPFC mQTLs near *APOC2*, with 2 implicated CpG sites also associating with CAA and APOC2 expression, yet also noting that *APOC2* expression was not significantly associated with the severity of CAA in ROSMAP (P=0.089). Across these observations and incongruent eQTL results, they reasonably concluded that *APOC2* and related methylation may be the culprit in their CAA genetic signal.

**Table 1.**
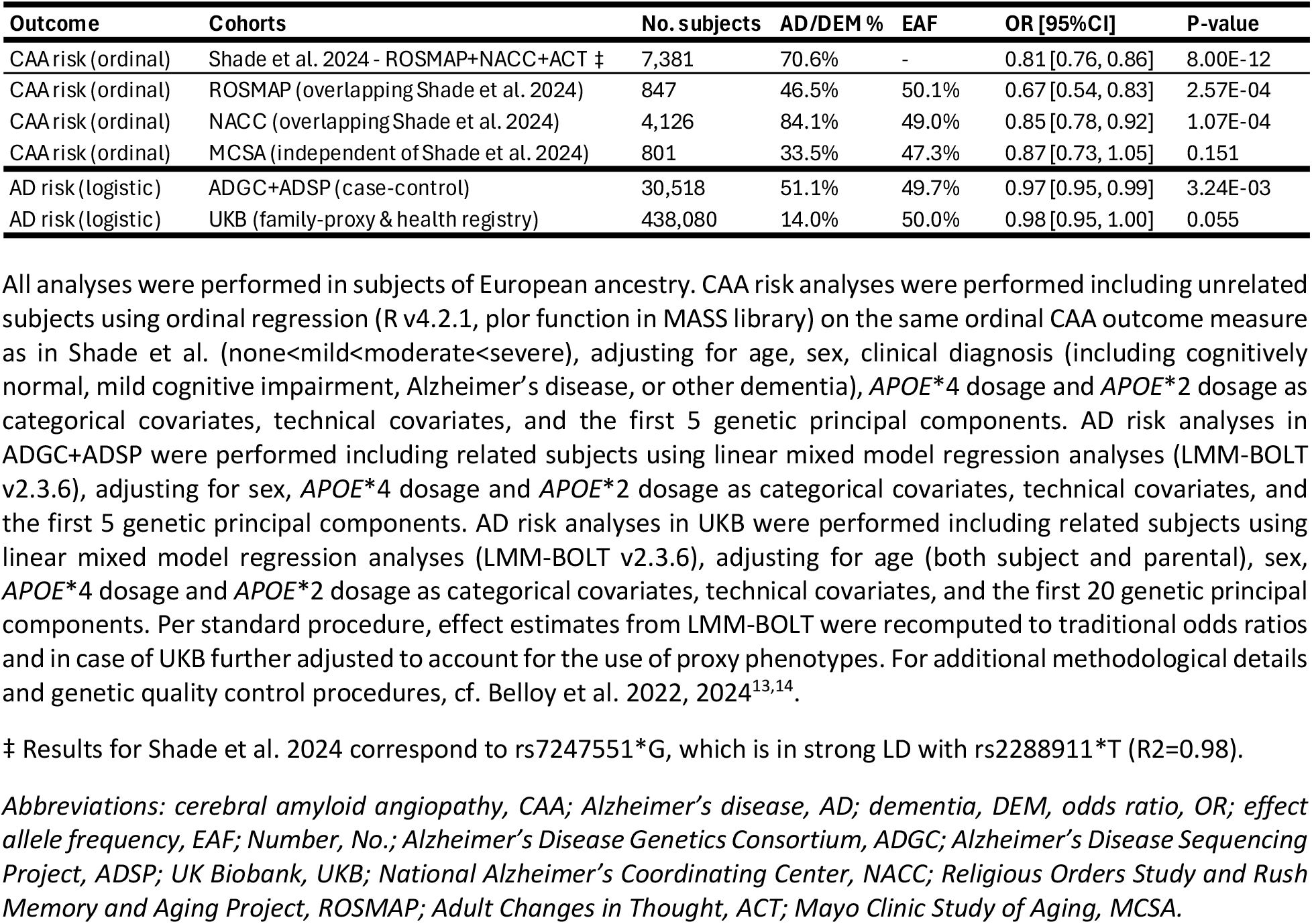
Association of rs2288911*T with cerebral amyloid angiopathy and Alzheimer’s disease.

However, earlier this year, Fujita et al. presented a large single-nucleus RNAseq eQTL resource (n=424) in ROSMAP DLPFC (largely overlapping the bulk DLPFC eQTL data evaluated by Shade et al.)^11^. Strikingly, they found a microglia-specific eQTL overlapping the locus identified by Shade et al. Their top variant, rs2288911, is in strong linkage disequilibrium (LD) with rs7247551 (R2=0.98). rs2288911 lies upstream, 5Kb closer to *APOE*, within a site with marks for microglia-specific active enhancers (H3K27ac) and promoters (H3K4me3), as well as evidence for chromatin interactions with *APOE* (PLAC-seq) (cf. Fig.4d in Fujita et al.). They also evaluated associations of rs2288911 with different traits in ROSMAP, highlighting an association with CAA independent of *APOE*\*4 (P=9.9e-6). The rs2288911*T allele, correlated with the protective rs7247551*G allele in Shade et al., was associated with decreased *APOE* expression in microglia and decreased CAA risk.

## Results & Discussion

A new inspection of the microglia eQTL data from Fujita et al. illustrates that rs2288911 is an eQTL for *APOE* but not *APOC2* (**Fig.1A**). This observation replicates in a prior, independent microglia eQTL meta-analysis (n=400) by Kosoy et al. (**Fig.1B**)^12^, further corroborating that *APOE* expression in microglia, rather than *APOC2*, is the likely mechanism behind this CAA genetic signal. However, it should be noted that rs2288911 appears to be an equally significant eQTL for *APOC1* (**Fig.S1**), which is the closest downstream gene from *APOE*. Co-regulation of nearby genes is common, and there is to our knowledge no literature implicating *APOC1* in amyloid or CAA pathology, such that *APOE* remains the most likely candidate gene, although future work should consider a potential role of *APOC1*.

**Figure 1.**
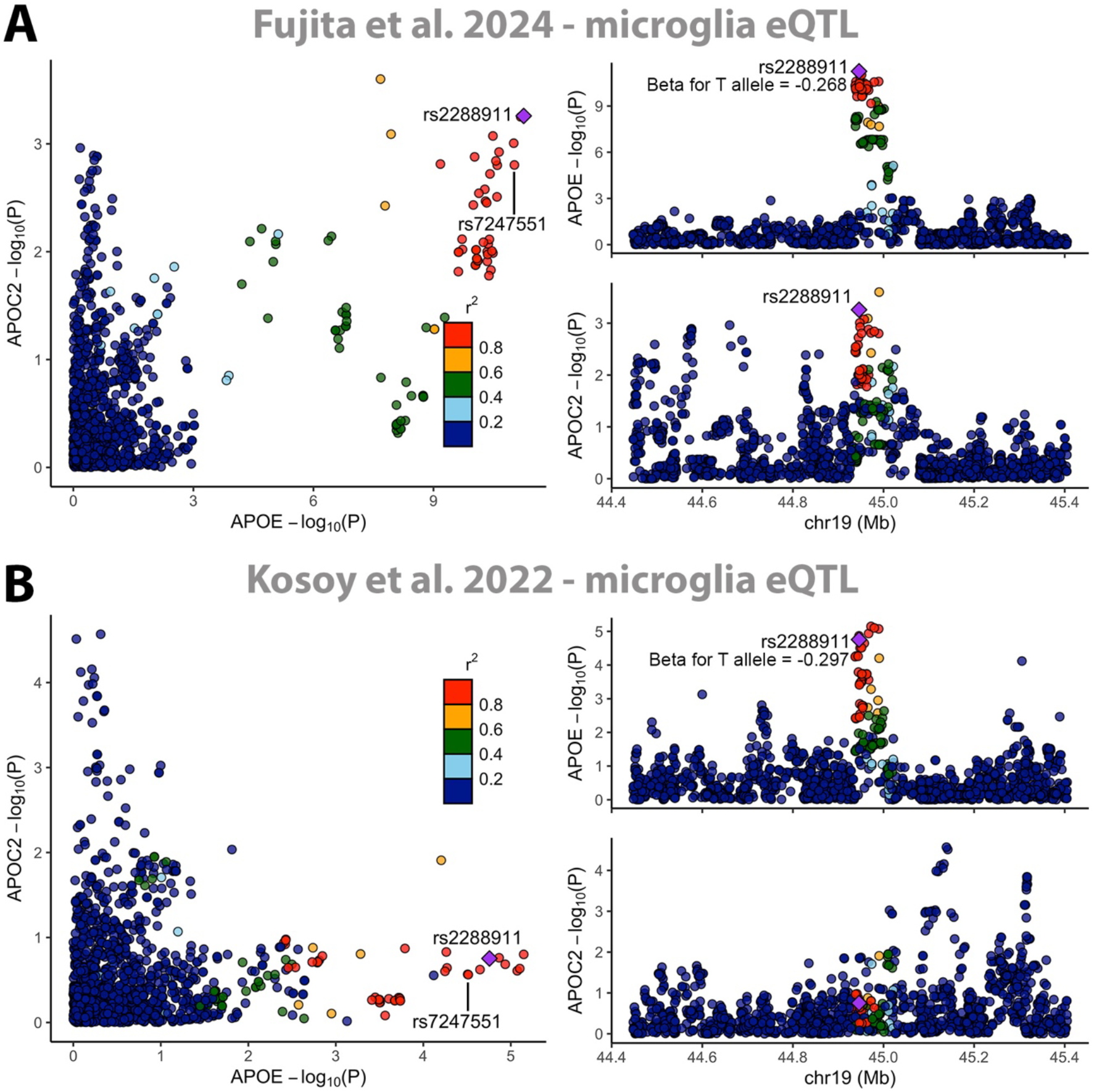
Rs2288911 and rs7247551 are microglia expression quantitative trait loci for *APOE*, not *APOC2*. Locus comparison (left) and locus zoom (right) plots indicate that the two SNPs are microglia eQTLs for *APOE* and *APOC2* in two independent datasets: **(A)** Fujita et al. 2024 (n=424) and **(B)** Kosoy et al. 2022 (n=400). Color coding of variants/dots indicates LD (R2) with rs2288911, marked by purple diamonds (R v4.2.1, locuscomparer library). Plots only display variants present in both datasets (93% overlap within depicted 1Mb range). rs2288911 was identified in Fujita et al. as the lead microglia-specific eQTL for *APOE* and is in strong LD with rs7247551 identified by Shade et al. (R2=0.98), who related its effect to *APOC2* and local methylation. rs2288911 shows only a nominal association with microglial *APOC2* expression in (A) and none in (B), indicating it is not an eQTL for *APOC2*. Conversely, it is strongly associated with *APOE* expression in both datasets with consistent beta coefficients that suggest close to 30% reduced microglia *APOE* expression for a single T allele. *APOE* eQTL signals across (A) and (B) colocalized with PP4=0.94 (R v4.2.1, coloc library). *Abbreviations: expression quantitative trait locus, eQTL; Linkage disequilibrium, LD; Colocalization poster probability for hypothesis 4: association with both traits and a shared variant, PP4*.

Unbeknownst to Shade et al., the earlier Fujita et al. study inspired us to look at the association of rs2288911 with CAA and AD risk in large genetic datasets from the Alzheimer’s Disease Genetics Consortium (ADGC), Alzheimer’s Disease Sequencing Project (ADSP), and UK Biobank (UKB). We leveraged our established pipelines and state-of-the-art *APOE* genotype quality control to ensure the most accurate assessment of genetic associations conditioned on *APOE*\*4 and *APOE*\*2^13,14^. We reproduced the Fujita et al. rs2288911*T-CAA association in ROSMAP and additionally replicated this in NACC data overlapping ADGC and ADSP, effectively reproducing the CAA finding in Shade et al. (**Table.1**). We further added an additional replication attempt in the population-based Mayo Clinic Study of Aging (MCSA, n=801)^15^, where the effect direction was consistent but diminished and non-significant (**Table.1**). The MCSA primarily follows cognitively unimpaired individuals or those with mild cognitive impairment. Notably, Shade et al. also performed replication analyses in the MAYO brain bank, specifically MC-CAA (n=815)^3^, and obtained a significant replication (P=0.0012) using an alternative CAA phenotype. While some overlap in sample is present, MC-CAA includes only participants that met neuropathological criteria for AD (NINCDS-ADRDA), with Braak stage>3, while MCSA contained fewer advanced cases. However, sensitivity analyses across NACC and ROSMAP did not indicate a stronger effect size at higher Braak stages, marking some unexplained variability in effect estimates (Braak≤3, OR=0.72, 95%CI=[0.60,0.85], P=2.0e-4, n=1,165; Braak>3, OR=0.85, 95%CI=[0.79,0.94], P=7.2e-4, n=3,735). Finally, we assessed the association of rs2288911*T with AD risk (**Table.1**). It’s crucial to appreciate here that even the slightest LD of variants at the *APOE* locus with *APOE*\*4 or *APOE*\*2 may inflate association results, which is additionally relevant as AD-specific *APOE*-haplotypes may exist. We thus verified that *APOE*\*4 (rs429358) and *APOE*\*2 (rs7412) have very low (though still detectable) LD with rs2288911 in unrelated case-control samples from ADGC and ADSP (respectively R2=0.00223 and R2=0.00249). We then observed that rs2288911*T is associated with reduced AD risk even after conditioning on *APOE*\*4 and *APOE*\*2 (**Table.1**). This is the first analysis at this scale to provide association results for rs2288911 with AD risk conditioned on *APOE*\*4 and *APOE*\*2. The effect of rs2288911 on AD risk is consistent with the shared genetic contributors to AD and CAA. The weaker signal in AD compared to CAA supports the consensus that CAA and AD are only partially genetically correlated and that there is still important biological distance between the two diseases.

## Conclusion

In summary, Shade et al., Fujita et al., and our group, provide compelling evidence for the association of rs2288911*T or rs7247551*G at the *APOE* locus with lower CAA risk, independent of *APOE*\*2 and *APOE*\*4. Seeking to reconcile findings across these studies, we suggest the most promising mechanism is that the causal variant reduces *APOE* expression in microglia specifically. Our additional analyses confirm the variant is also associated with AD risk in common AD GWAS cohorts, independent of *APOE*\*2 and *APOE*\*4.

Lastly, we suggest that further exploration of this CAA association, as well as additional replication efforts, will be relevant to confirm its robustness. In conclusion, this genetic signal for CAA at the *APOE* locus and the proposed mechanism represent important targets for further experimental validation, which is bound to generate support for the clinical relevance of lowering APOE to protect against CAA and AD.

## Methods

All genetic association analyses were performed in subjects of European ancestry. CAA risk analyses were performed including unrelated subjects using ordinal regression on the same ordinal CAA outcome measure as in Shade et al. (none<mild<moderate<severe), adjusting for age, sex, clinical diagnosis (including cognitively normal, mild cognitive impairment, Alzheimer’s disease, or other dementia), *APOE*\*4 dosage and *APOE*\*2 dosage as categorical covariates, technical covariates, and the first 5 genetic principal components. AD risk analyses in the Alzheimer’s Disease Genetics Consortium (ADGC) and Alzheimer’s Disease Sequencing Project (ADSP) data were performed including related subjects using linear mixed model regression analyses (LMM-BOLT), adjusting for sex, *APOE*\*4 dosage and *APOE*\*2 dosage as categorical covariates, technical covariates, and the first 5 genetic principal components (PCs). AD risk analyses in UK Biobank (UKB) were performed including related subjects using linear mixed model regression analyses (LMM-BOLT), adjusting for age (both subject and parental), sex, *APOE*\*4 dosage and *APOE*\*2 dosage as categorical covariates, technical covariates, and the first 20 genetic PCs.

Expression quantitative trait locus (eQTL) colocalization analyses for genes at the *APOE* locus were assessed from (1) Fujita et al. 2024 for single nucleus (microglia + 6 additional cell types) data from dorsolateral prefrontal cortex in ROSMAP (N=424) and (2) Kosoy et al. 2022 for single cell (microglia) data meta-analyzed across multiple brain regions and studies (N=400). QTL genetic colocalization analyses were conducted using the coloc library, R v4.2.1.

## Data Availability

Data used in the CAA and AD risk analyses are available upon application to:

- dbGaP (https://www.ncbi.nlm.nih.gov/gap/)
- NIAGADS (https://www.niagads.org/)
- LONI (https://ida.loni.usc.edu/)
- AMP-AD knowledge portal / Synapse (https://www.synapse.org/)
- Rush (https://www.radc.rush.edu/)
- NACC (https://naccdata.org/)
- UKB (https://www.ukbiobank.ac.uk/)
- MCSA (https://www.mayo.edu/research/centers-programs/alzheimers-disease-research-center/data-requests)

The specific data repositories and identifiers for ADGC and ADSP data are indicated in **Table.S1** of the supplement.

Microglia eQTL datasets are available from:

- Fujita et al. 2024 - AMP-AD knowledge portal / Synapse (https://www.synapse.org/; identifier: syn52335732)
- Kosoy et al. 2022 - AMP-AD knowledge portal / Synapse (https://www.synapse.org/; identifier: syn26207321)

## Code Availability

All presented analyses made use of standard functionalities available in R v4.2.1 (libraries: “plor”, “coloc”, and “locuscomparer”) and LMM-BOLT v2.3.6. Processing of genetic and phenotypic data is described in Belloy et al. 2022, 2024^13,14^.

## Consent for publication

Not applicable.

## Competing/Conflicting interests

The authors declare no competing or conflicting interests.

## Ethics oversight

The current study protocol was granted an exemption by the Stanford Institutional Review Board because the analyses were carried out on “de-identified, off-the-shelf” data; therefore, additional informed consent was not required.

## Funding/Support

Funding for this study was provided by the NIH (R00AG075238, M.E.B; AG060747 and AG047366, M.D.G; AG006786, J.G-R.)

## Role of Funder/Sponsor

The funding organizations and sponsors had no role in the design and conduct of the study; collection, management, analysis, and interpretation of the data; preparation, review, or approval of the manuscript; and decision to submit the manuscript for publication.

## Authors’ contributions

M.E.B. and M.D.G. had full access to all the data in the study and take responsibility for the integrity of the data and the accuracy of the data analysis. M.E.B. performed data acquisition and analyses, designed analyses, designed study, wrote paper, and obtained funding. J.G-R. was involved in data, funding, and resource acquisition. M.D.G designed study, designed analyses, supervised work, and obtained funding. All authors contributed to critical revision of the manuscript.

## Acknowledgements

Data for this study were prepared, archived, and distributed by the National Institute on Aging Alzheimer’s Disease Data Storage Site (NIAGADS) at the University of Pennsylvania (U24-AG041689), funded by the National Institute on Aging. The contents of this article do not represent the views of the National Institutes of Health, the U.S. Department of Veterans Affairs, or the United States Government. Additional data specific acknowledgements are provided in the supplementary material.

## Notes

### Competing Interest Statement

The authors have declared no competing interest.

### Author Declarations

The current study protocol was granted an exemption by the Stanford Institutional Review Board because the analyses were carried out on de-identified, off-the-shelf data; therefore, additional informed consent was not required.

